# Antigenic evolution of SARS-CoV-2 in immunocompromised hosts

**DOI:** 10.1101/2022.01.13.22269154

**Authors:** Cameron A. Smith, Ben Ashby

**Affiliations:** Department of Mathematical Sciences, University of Bath, Bath, BA2 7AY, UK; Milner Centre for Evolution, University of Bath, Bath, BA2 7AY, UK; Department of Mathematics, Simon Fraser University, Burnaby, BC, V5A 1S6, Canada

**Keywords:** Antigenic evolution, SARS-CoV-2, COVID-19, variant, immunocompromised, epistasis

## Abstract

Prolonged infections of immunocompromised individuals have been proposed as a crucial source of new variants of SARS-CoV-2 during the COVID-19 pandemic. In principle, sustained within-host antigenic evolution in immunocompromised hosts could allow novel immune escape variants to emerge more rapidly, but little is known about how and when immunocompromised hosts play a critical role in pathogen evolution. Here, we use a simple mathematical model to understand the effects of immunocompromised hosts on the emergence of immune escape variants in the presence and absence of epistasis. We show that when the pathogen does not have to cross a fitness valley for immune escape to occur (no epistasis), immunocompromised individuals have no qualitative effect on antigenic evolution (although they may accelerate immune escape if within-host evolutionary dynamics are faster in immunocompromised individuals). But if a fitness valley exists between immune escape variants at the between-host level (epistasis), then persistent infections of immunocompromised individuals allow mutations to accumulate, therefore facilitating rather than simply speeding up antigenic evolution. Our results suggest that better genomic surveillance of infected immunocompromised individuals and better global health equality, including improving access to vaccines and treatments for individuals who are immunocompromised (especially in lower- and middle-income countries), may be crucial to preventing the emergence of future immune escape variants of SARS-CoV-2.

**Lay Summary:** We study the role that immunocompromised individuals may play in the evolution of novel variants of the coronavirus responsible for the COVID-19 pandemic. We show that immunocompromised hosts can be crucial for the evolution of immune escape variants. Targeted treatment and surveillance may therefore prevent the emergence of new variants.

## Introduction

Understanding how and when variants of SARS-CoV-2, the causative agent of COVID-19, are likely to evolve is key to managing the future of the pandemic. Multiple variants of concern have evolved since the start of the pandemic, with higher transmissibility evolving on at least two occasions, by the Alpha (B.1.1.7) variant (relative to the wildtype) [1], and by the Delta (B.1.617.2) variant (relative to Alpha) [2,3], with the latter becoming the globally dominant strain in 2021 [4]. Other variants such as Beta (B.1.351) and Omicron (B.1.1.529) have additionally shown evidence of immune escape, indicating antigenic evolution [5–7] (Omicron has also been linked with an increase in transmission [8,9]). With increasing numbers of people acquiring immunity to SARS-CoV-2, either through infection or vaccination, we should expect a shift towards antigenic evolution rather than higher intrinsic transmissibility or greater virulence as the primary driver of new variants of concern [10]. The extent to which SARS-CoV-2 may evolve antigenically in future, thereby allowing it to evade host immunity partially or fully, is currently unknown. However, the emergence and rapid spread of Omicron towards the end of 2021 has demonstrated that antigenic evolution is both possible and under strong selection. The unusual nature of Omicron (possessing a large number of mutations in the spike protein but only distantly related to the dominant variant at the time, Delta [11]) has led to speculation that it underwent long-term within-host evolution in an immunocompromised individual who was unable to clear the infection [12]. We explore this hypothesis using a simple mathematical model to understand the potential importance of immunocompromised individuals for the antigenic evolution of SARS-CoV-2.

A fundamental tenet of evolutionary epidemiology is that the rate of antigenic evolution depends on a balance between immune pressure and mutation supply [13–15]. The greater the proportion of the population that is immune, the greater the strength of selection for immune escape but mutation supply is constrained as few hosts can be infected. Conversely, if many hosts are susceptible to infection, then mutation supply may be plentiful but selection for immune escape is relatively weak. Hence, the rate of antigenic evolution should be maximised at an intermediate level of immune pressure, whereby moderate pathogen prevalence leads to a plentiful supply of mutations for selection to act upon, and the strength of selection for immune escape is reasonably strong.

Rapid deployment of vaccinations against SARS-CoV-2 combined with the relaxation of non-pharmaceutical interventions in many countries led to both strong immune pressure and high numbers of infections in the latter half of 2021. For example, by the end of November 2021 the UK had fully vaccinated 68% of the population while still experiencing over 620 confirmed cases per million (approximately 70% of the previous peak in January 2021) [16]. At the time, the Delta variant was dominant globally and accounted for over 99% of infections in the UK [16]. Yet, despite apparently favourable evolutionary conditions for immune escape there were no indications of the Delta variant exhibiting antigenic evolution in the UK or elsewhere. This may indicate that closely related immune escape variants were suppressed, perhaps by transiently boosted innate, B, and T cell responses, or due to epistasis (e.g. less transmissible). Instead, the initial BA.1 sublineage of the Omicron variant, first detected in South Africa and reported to the World Health Organization on November 24, 2021 [11], was able to substantially escape host immunity and evolved from a distant clade. This variant contains 30 mutations to the spike protein (used for binding to host cell receptors) and has been shown to evade over 85% of neutralizing antibodies [7]. Relative to Delta, it exhibits substantially lower vaccine effectiveness [17] and is estimated to be over five times as likely to lead to reinfection [6]. The BA.1 sublineage of Omicron became the dominant variant in the UK within a month and replaced Delta in many countries in early 2022 [16], with the BA.2 sublineage later replacing BA.1 [18].

The BA.1 sublineage of Omicron confirms that substantial immune escape is not only possible for SARS-CoV-2 but also that selection for immune escape towards the end of 2021 was very strong. According to the conceptual model of antigenic evolution as a balance between immune pressure and mutation supply [13], this suggests that the lack of adaptation to evade host immunity by the Delta lineage was simply due to insufficient mutation supply. However, this is difficult to reconcile with the high number of cases at the time, implying mutation supply was plentiful. Furthermore, if mutation supply was the key constraint, how did an immune escape variant appear from an obscure clade that was responsible for few infections?

Several hypotheses have been proposed for the sudden emergence of the Omicron variant from a distant clade. One possibility is that omicron evolved in an animal host following infection by a human, and then jumped back into the human population. Alternatively, it could have evolved in a remote population without being detected until it began to spread more widely in late 2021. However, neither of these explanations are especially convincing. Evolution in an animal host would have not only required two jumps across the human-animal species barrier, but also selection in the animal host would have had to correspond to increased fitness in the human population through immune escape. It is more plausible that Omicron was able to substantially escape immunity in humans because it had experienced selection for immune escape in humans. Similarly, evolution in a remote population does not appear to be plausible as it fails to explain why a similar array of mutations were not seen in regions where mutation supply was significantly higher (due to more infections) and immune pressure was strong due to vaccine and naturally-acquired immunity.

A more promising hypothesis is that the Omicron variant arose due to long-term within-host evolution in an immunocompromised individual, who was most likely infected between March and August 2021 [11]. While an immunocompetent individual would be expected to clear infection after a relatively short period, an immunocompromised person may fail to fully clear the infection, allowing the virus to coevolve with the immune system [19]. Indeed, longitudinal sequencing from an immunocompromised patient who was infected for over 150 days with SARS-CoV-2 revealed rapid accumulation of mutations [20]. These mutations appeared to be adaptive at the within-host level due to their concentration in the spike protein, with several common to other variants of concern. Similar results have been observed in other patients with long-term infections of SARS-CoV-2 [21,22], including those who have been treated with convalescent plasma, indicating antigenic evolution within the host [23] (although some immunocompromised individuals show little to no within-host evolution of SARS-CoV-2 [24]). A study of infection in immunocompromised individuals has found that mutations accumulate in the spike gene receptor binding domain and N-terminal domains, associated with immune escape and viral packaging [25]. Furthermore, a recent study has concluded that the large number of mutations which were associated with the Alpha variant likely occurred in an immunocompromised individual [26].

It is currently unclear how important immunocompromised individuals are for the antigenic evolution of SARS-CoV-2, or for pathogen evolution more generally. Do infections of immunocompromised individuals simply accelerate antigenic evolution or do they play a key role in facilitating immune escape? In the first case, infection of immunocompromised individuals speeds up antigenic evolution due to a faster rate of adaptation within these hosts, leading to the emergence of new immune escape variants on shorter timescales than would be possible in an immunocompetent population. Such a scenario would suggest that although immunocompromised individuals might speed up antigenic evolution, they are not essential for it to occur. In the second case, long-term infections of immunocompromised individuals allow the virus to accumulate mutations that are advantageous (or neutral) at the within-host level but may be disadvantageous (or neutral) at the between-host level. If there is epistasis between mutations at the between host level (i.e. fitness depends on the context of which other mutations are present), as indicated by recent experiments [27], then sustained adaptation within immunocompromised individuals may allow the virus to traverse valleys in the fitness landscape, which would otherwise be very difficult to cross, to reach another peak. The second scenario would therefore suggest that long-term infections in immunocompromised individuals play a disproportionate role in the antigenic evolution of pathogens such as SARS-CoV-2.

Here, we analyse a simple phenomenological model to explore the potential importance of immunocompromised hosts for the antigenic evolution of SARS-CoV-2. We show that in the absence of epistasis, antigenic evolution readily occurs regardless of the frequency of immunocompromised individuals in the population. If epistasis is present, however, such that the virus must traverse a fitness valley at the between-host level to escape host immunity, then immunocompromised hosts are crucial for antigenic evolution to occur. These patterns are robust irrespective of whether within-host evolutionary dynamics are faster in immunocompromised individuals and for a wide range of parameters affecting cross-immunity, the strength of epistasis, the proportion of the population that is immunocompromised and their duration of infection relative to immunocompetent hosts.

### Model description

We adapt the model of antigenic evolution presented by Gog and Grenfell [28] to incorporate immunocompromised individuals and epistasis. The model assumes that there are *n* = 30 variants equally spaced in a line, with adjacent variants differing by a single mutation. Hosts are classed as either entirely susceptible to a variant, or entirely immune to it. Cross-immunity between variants is therefore ‘polarising’, which means that when an individual is infected by variant *i*, a proportion *σ*_*ij*_ of those currently susceptible to variant *j* become fully immune to it for life (no waning immunity) and a proportion 1 − *σ*_*ij*_ remain fully susceptible to variant *j*. This assumption greatly reduces the complexity of the model as it means we do not need to track all infection histories, which would require at least (2 + *n*)2^*n*^ ≈ 137 billion classes with *n* = 30. The strength of cross-immunity between variants *i* and *j* is given by

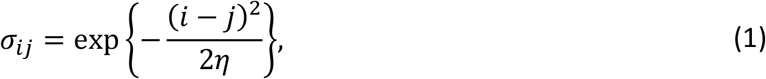

where *η* > 0 controls the breadth of cross-immunity (large values of *η* give broad cross-immunity between distant variants, whereas small values of *η* limit cross-immunity to closely related variants; Fig. 1a). We assume that the population is large, well-mixed, and of constant size (*N* = 10^7^), with a proportion *p* of individuals who are immunocompromised (only able to produce a weak immune response; subscript *C*) and a proportion 1 − *p* who are immunocompetent (able to produce a normal or “healthy” immune response; subscript *H*). For simplicity, we ignore host demographics (births and deaths) and mortality from infection, as we are only interested in the antigenic evolution of the virus over a relatively short timescale.

**Figure 1:**
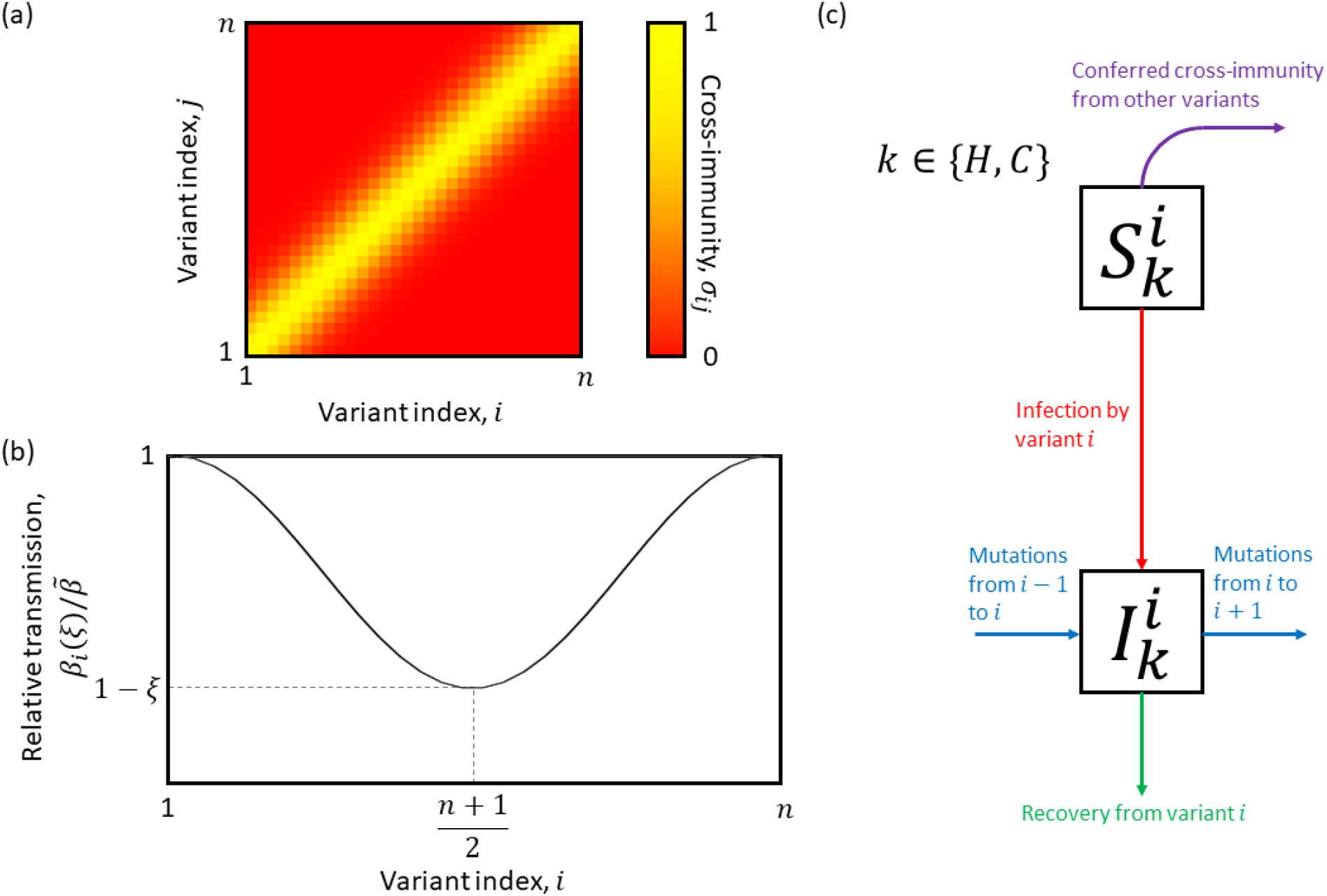
Population-level model. (a) Cross-immunity,σ_ij_, for variants i and j, with lighter colours corresponding to greater cross-immunity. (b) Illustration of the normalised transmission rate for each variant, showing a fitness valley. (c) Model schematic.

Let 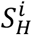 (respectively 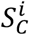) be the proportion of the population that is immunocompetent (resp. immunocompromised) and susceptible to variant *i* ∈ {1, …, *n*}, and 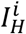 (resp. 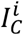) be the proportion of the population that is immunocompetent (resp. immunocompromised) and infected with variant *i*. To incorporate a fitness valley at the between-host level, we assume that the transmission rate of variant *i* is given by

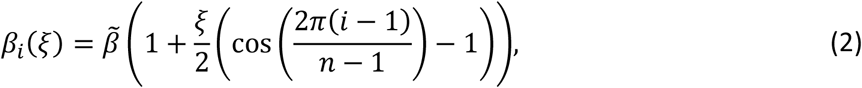

where 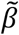 is the maximum transmission rate and *ξ* controls the strength of epistasis (Fig. 1b). Preliminary analysis revealed that other functional forms with qualitatively similar properties produce results consistent with those presented below. When *ξ* = 0, there is no epistasis as 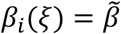 for all variants. When 0 < *ξ* < 1, epistasis reduces the transmission rate for variants intermediate between 1 and *n*, reaching a minimum of *β*_*i*_(*ξ*) = 1 − *ξ*, with 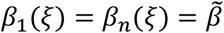 for all *ξ* (Fig. 1b).

Healthy and immunocompromised individuals are identical in our model except for their infectious periods and rates of within-host antigenic evolution. The infectious period for immunocompromised individuals, 1/*γ*_*C*_ = 140 days, is assumed to be 20 times longer than that for healthy individuals, 1/*γ*_*H*_ = 7 days. These values are chosen to be illustrative and reasonable parameter variation does not qualitatively affect our results. The rate of within-host antigenic evolution (i.e. the per-capita transition rate between adjacent variants in the antigenic space) is governed by parameters *μ*_*H*_ and *μ*_*C*_ in healthy and immunocompromised individuals, respectively. We assume that the virus mutates at a constant rate, leading to a constant rate of antigenic evolution for a given host type. While our primary model implicitly captures a simplified version of within-host evolutionary dynamics by assuming a constant rate of antigenic evolution per host type, we justify this assumption by exploring a separate within-host only model in the Appendix, which demonstrates that a constant rate of antigenic evolution is a reasonable approximation. In our primary model, we either set *μ*_*H*_ = *μ*_*C*_ so that the rate of within-host antigenic evolution is the same in healthy and immunocompromised hosts, or set *μ*_*H*_ < *μ*_*C*_ to investigate the impact of a faster rate of within-host antigenic evolution in immunocompromised individuals. The rate of antigenic evolution may differ between host types due to differences in the viral population size within a host or the strength of selection for immune escape.

To allow for random mutations, we simulate our model using the stochastic *τ*-leaping method [29] for the underlying ordinary differential equations (ODEs)

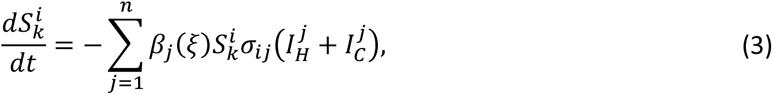

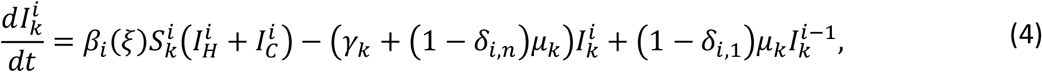

where *δ*_*i,j*_ is the Kronecker delta, which takes the value 1 if *i* = *j* and is 0 otherwise. A schematic for this system can be found in Fig. 1c.

We run 10 simulations for each parameter combination up to *t*_*max*_ = 1460 time steps (days), as preliminary analysis revealed that either antigenic evolution reaches the boundary of antigenic space within this timeframe, or the infection is driven extinct. Note however, that this duration is arbitrary and varies inversely with *μ*_*C*_ and *μ*_*H*_. We say that a variant is ‘observed’ if it exceeds a threshold of 0.01. In each simulation, we summarise the dynamics by measuring the total number of variants observed and the maximum distance in antigenic space between observed variants.

## Results

We wish to establish when antigenic evolution proceeds as a gradual diffusion through antigenic space, or in large jumps. We therefore focus our analysis on the strength of epistasis on transmissibility *ξ*, the strength of cross-immunity *η*, the proportion of the population that is immunocompromised *p*, the relative rate of adaptation (antigenic evolution) in immunocompromised hosts *μ*_*C*_/*μ*_*H*_ and the relative infectious period *γ*_*H*_/*γ*_*C*_. In the absence of epistasis (or when epistasis is sufficiently weak), the virus diffuses gradually through antigenic space (Figs. 2a and 2c). As the host population accumulates immunity to the current dominant variant, selection favours the next variant in line that can substantially escape immunity, leading to successive epidemic waves at regular intervals. This occurs regardless of whether within-host evolution is assumed to be faster in immunocompromised individuals (Fig. 2e).

**Figure 2:**
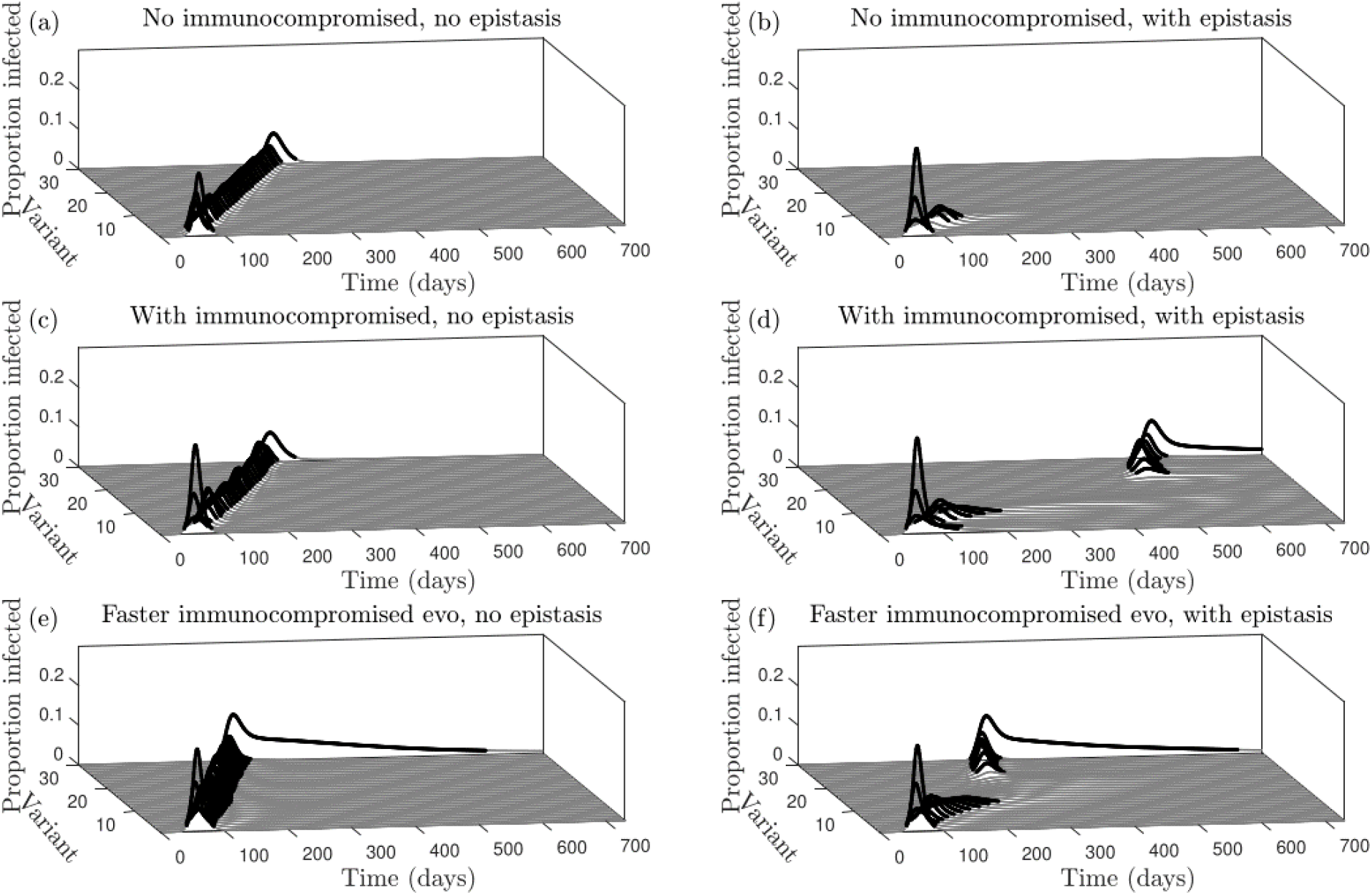
Antigenic evolution with or without immunocompromised individuals and epistasis. *(a) No epistasis in an entirely immunocompetent population* (*p* = 0, *ξ* = 0). *(b) Strong epistasis in an entirely immunocompetent population* (*p* = 0, *ξ* = 0.8). *(c) No epistasis and a small immunocompromised subpopulation* (*p* = 0.05, *ξ* = 0). *(d) Strong epistasis and a small immunocompromised subpopulation* (*p* = 0.05, *ξ* = 0.8). *(e) No epistasis and a small immunocompromised subpopulation with faster within-host evolution in immunocompromised individuals (p* = 0.05, *ξ* = 0.8, *μ*_*C*_ = 5*μ*_*H*_*). (f) Strong epistasis and a small immunocompromised subpopulation with faster within-host evolution in immunocompromised individuals (p* = 0.05, *ξ* = 0.8, *μ*_*C*_ = 5*μ*_*H*_*). All other parameter values given in Table A3. Dynamics are shown for a single simulation*.

When epistasis is sufficiently strong, however, the proportion of the population that is immunocompromised plays a crucial role in antigenic evolution (Figs. 2b, d, f). If very few individuals are immunocompromised, the epidemic quickly burns out with little antigenic evolution, as the virus is unable to cross the fitness valley caused by epistasis at the between-host level (Fig. 2b). But if a sufficient proportion of the population is immunocompromised, then the virus can cross this fitness valley due to within-host evolution in this subpopulation (Fig. 2d). Immunocompromised hosts experience longer infections, on average, which allows the virus to accumulate mutations and cross the fitness valley. When the virus has acquired enough mutations in the immunocompromised such that between-host transmissibility is restored to a sufficiently high level, it is able to spread in the rest of the host population. Again, this process is sped up if the within-host evolutionary dynamics are assumed to be faster in immunocompromised individuals, but the qualitative dynamics are unchanged (Fig. 2f).

Our results are qualitatively robust to variation in key model parameters, although our sensitivity analysis reveals two notable interactions (Fig. 3). When varying the strength of epistasis and the extent of cross-immunity between variants, we find that, intuitively, immunocompromised individuals are especially important for traversing the fitness valley if epistasis is stronger or if cross-immunity is broader (Fig. 3a). This is because stronger epistasis makes the fitness valley deeper and broader cross-immunity reduces the pool of susceptible hosts across a wider range of variants. However, if epistasis is sufficiently strong (around *ξ* = 0.8 in Fig. 3a) a large jump in antigenic space to a distant variant occurs regardless of the strength of cross-immunity. Our sensitivity analysis also reveals that as the proportion of the population that is immunocompromised decreases, a jump in antigenic space becomes less likely and requires a longer relative infectious period in immunocompromised hosts (Fig. 3b). This suggests that better treatment of immunocompromised hosts (to reduce the average duration of infection), improved genomic surveillance of these hosts (to identify novel variants of concern), and better prevention and treatment of pre-existing conditions (to reduce the proportion of the population that is immunocompromised) may greatly reduce the likelihood of new variants emerging at distant fitness peaks.

**Figure 3:**
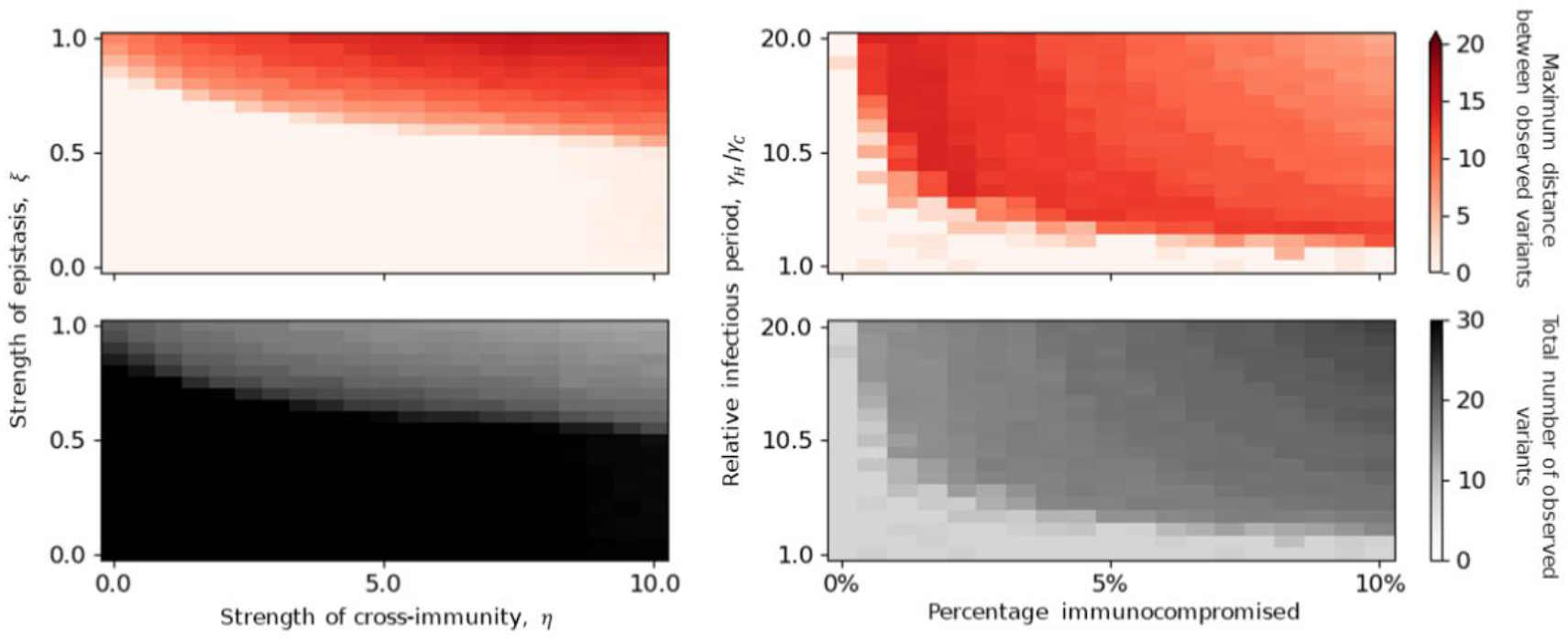
Sensitivity analysis. *Top row: maximum distance between observed variants (darker shading indicates larger jumps in antigenic space); bottom row: total number of variants observed. (a) Varying the strength of cross-immunity* (*η*) *and epistasis* (*ξ*) *when 5% of the population is immunocompromised (p=0*.*05). (b) Varying the percentage of the population that is immunocompromised and the relative recovery periods (with η* = 5 *and ξ* = 0.8*). All other parameters as in Table A3. All datapoints are averaged over 10 simulations*.

## Discussion

The presence of immunocompromised individuals has been suggested as an important driver behind not only the emergence of the Omicron variant of SARS-CoV-2, but also other variants of concern, including Alpha and Delta [20]. Using a simple model of antigenic evolution, we have shown that prolonged infections of immunocompromised individuals allow pathogens to accumulate sufficient mutations to overcome epistasis at the between-host level, facilitating the emergence of novel immune escape variants. Our model was motivated by the sudden emergence of the Omicron variant from a distant clade to the dominant variant at the time, coupled with longitudinal sequencing from immunocompromised patients that indicate rapid within-host evolution [18–20]. Given relatively high levels of infection (and hence mutation supply; Fig. 4) combined with rapidly increasing immune pressure in mid-to late-2021, conditions for the Delta lineage to exhibit antigenic evolution seemed to be favourable. Mutation supply or lack of immune pressure therefore do not appear to have been the fundamental constraint for the lack of antigenic evolution by the Delta variant, which suggests that either epistasis or transiently-boosted innate immunity constrained immune escape. Indeed, our model suggests that novel immune escape variants readily evolve when epistasis is relatively weak. When epistasis is stronger, reducing transmissibility for variants between fitness peaks, we find that immunocompromised individuals may play a key role in antigenic evolution, effectively allowing the pathogen to traverse a fitness valley to reach a new peak. Note that while faster within-host adaptation in immunocompromised individuals speeds up the rate of antigenic evolution, unlike epistasis it does not qualitatively affect the outcome. Crucially, our results also suggest that improving treatment for those who are immunocompromised can greatly reduce the likelihood of new variants emerging.

**Figure 4:**
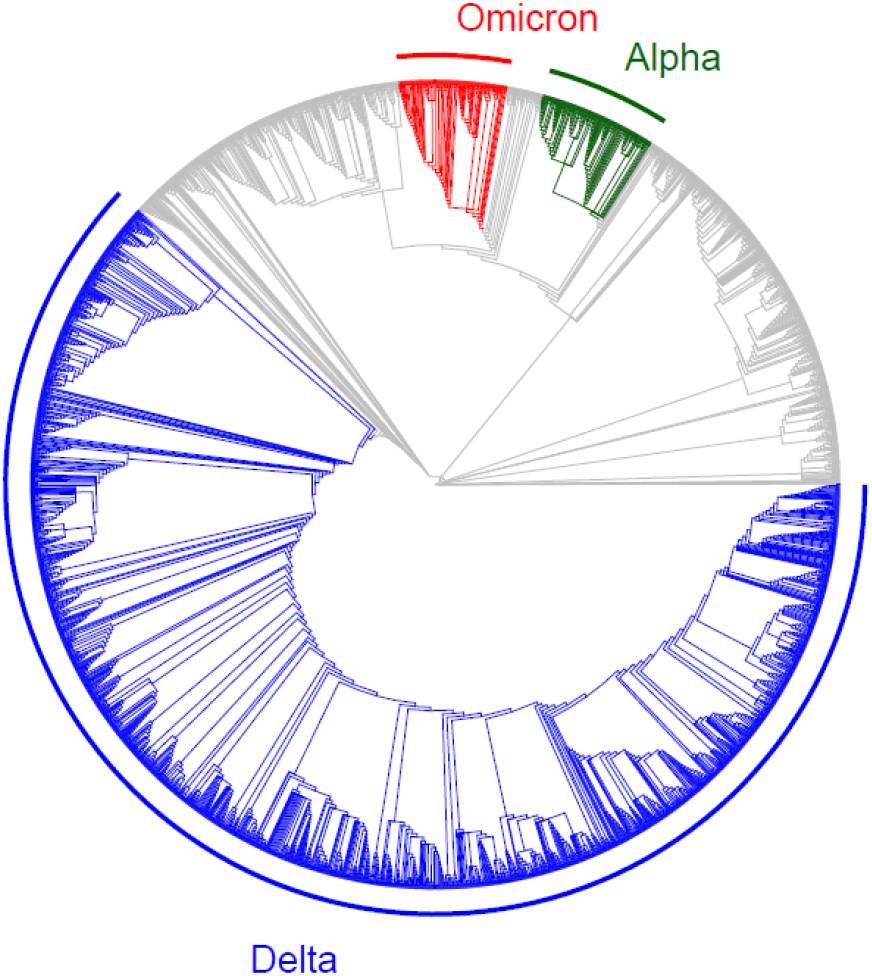
Phylogenetic tree for SARS-CoV-2 variants shortly after the emergence of Omicron. *Three variants of concern (Alpha, Delta and Omicron) are highlighted to illustrate that there had been high mutation supply for the Delta variant. Data downloaded from Nextstrain (nextstrain*.*org) on 08/02/2022* [18,30] *and provided by the Global Initiative for Sharing All Influenza Data (GISAID, gisaid*.*org)* [31–33]. *Data plotted using the ggtree software package in R* [34–36].

In real populations, individuals cannot simply be classed as either healthy or immunocompromised; they vary in the extent to which they are able to mount an immune response due to age, comorbidities, genetic or other environmental factors. Fitness landscapes for the pathogen may also differ between individuals and populations and real antigenic space is likely to be far more complex than our simplified one-dimensional space (note that multi-dimensional antigenic evolution can lead to more varied behaviour including branching and coalescence of phylogenetic pathways; see for example [37] and [38]). While our model does not capture the full complexity of antigenic evolution in real populations, it has important implications for our understanding of future immune escape variants of SARS-Cov-2, and for pathogen evolution more generally. Crucially, our model suggests that the lack of antigenic evolution by Delta followed by the emergence of Omicron is consistent with epistasis constraining immune escape in Delta, but this epistasis may have been overcome if immunocompromised individuals were infected for sufficiently long periods. Hence, rather than simply accelerating antigenic evolution, prolonged infections of immunocompromised individuals may have been critical for the evolution of Omicron. In the preprint of this manuscript, which was written shortly after the emergence of Omicron, we tentatively speculated that the lack of antigenic evolution by Delta suggested it may be difficult for SARS-CoV-2 to escape immunity through incremental mutations, and future variants may require multiple (epistatic) mutations to substantially escape host immunity. However, the subsequent emergence of immune evasion sub-variants of Omicron such as BA.4 and BA.5 suggests that either Delta was unusual in its limited scope for antigenic evolution and that other variants do not experience similar constraints, or that Delta may have eventually exhibited antigenic evolution if Omicron had not emerged. Regardless, our model suggests that immunocompromised individuals may remain a source of new variants that can substantially escape immunity. While not a focus of the current study, in principle prolonged infections of immunocompromised individuals could also facilitate the emergence and coexistence of multiple variants lacking cross-immunity, allowing the pathogen population to occupy different niches in a multidimensional antigenic space [39].

Our results agree with previous models which suggest that immunocompromised individuals are more likely to facilitate or accelerate within-host pathogen evolution, for example due to a longer average duration of infection or higher viral load [40,41]. However, while we find immunocompromised hosts to play a crucial role in pathogen evolution at the population level, other studies have concluded the opposite as these individuals only make up a small proportion of infections [40,41]. The reason for this discrepancy is likely due to contrasting assumptions regarding within-host fitness, immunity, and traits under selection. For example, van Egeren et al. [41] assume that a fitness valley exists at the within-host level with two or three mutations required to cross, whereas our model assumes that the fitness valley only exists at the between-host level (transmission) but may require many more mutations to traverse. If a fitness valley exists at the within-host level, then intuitively the importance of immunocompromised individuals for pathogen evolution will be lower. In general, there is no reason why the fitness landscape should have the same shape at the within- and between-host levels, as these are two very different environments with potentially contrasting selection pressures (e.g. UV exposure, temperature, interactions with the immune system). In general, one should not expect that a beneficial mutation in one context will necessarily be beneficial in another (“antagonistic pleiotropy”). Some mutations may therefore confer a fitness advantage at the within-host level, while being neutral or detrimental for transmission. For example, a mutation that increases the growth rate in the lower lung may be advantageous at the within-host level, but may lead to fewer transmission stages being produced, resulting in lower fitness at the between-host level. In SARS-CoV-2, the mutation D796H protects against neutralising antibodies but reduces infectivity, unless the mutation ΔH69/ΔV70 is also present [23]. It is therefore reasonable to expect differences in the fitness landscape at the within and between-host levels. In addition to the different assumptions about the fitness landscape, the model by van Egeren et al. [41] also focused on a static measure of relative fitness and did not consider antigenic evolution explicitly, whereas in our model the fitness of a particular variant depends on the level of immunity in the population, and so will vary over the course of the epidemic. Nevertheless, both models concur that longer duration infections, especially those of immunocompromised individuals, can play a disproportionate role in the evolution of novel variants, and are of particular concern for SARS-CoV-2 evolution.

We assumed that the rate of antigenic evolution during an infection was constant (but may vary by host type), which was motivated by the separate within-host model discussed in the Appendix. For immunocompetent hosts, who typically clear infection within two weeks [42], this means that there is relatively little time for new variants to emerge for onwards transmission, which slows down adaptation and can prevent epistatic mutations accumulating. But for immunocompromised hosts, who may experience much longer infections (upwards of 150 days [20]), the coevolutionary dynamics between the virus and the host immune system could allow many (potentially epistatic) mutations to accumulate. Interestingly, this hypothesis is consistent with previous theoretical [43] and experimental [44–46] studies showing that coevolution can both accelerate adaptation and allow a pathogen to cross fitness valleys caused by epistasis. For simplicity, our model assumed that the shape of the fitness landscape was similar in immunocompromised and immunocompetent hosts, preventing the pathogen from specialising on one type of host. Specialisation on immunocompromised individuals has been observed for other pathogens (e.g. *Pseudomonas aeruginosa* infections in cystic fibrosis patients [47]) and longitudinal studies of prolonged SARS-CoV-2 infections reveal the rapid accumulation of many mutations [20–24], but it is unclear if and when this leads to specialisation with a reduction in fitness on immunocompetent hosts.

While our model is informative, it does not capture the true complexity of antigenic space, the impact of vaccinations and non-pharmaceutical interventions, variation in disease outcomes, and the evolution of other disease characteristics such as transmissibility and virulence. This is by design so that our model requires as few assumptions as possible and so that the model can be adapted for other pathogens in future. We did not attempt to capture these effects, as our results are intended to be illustrative of the key roles that epistasis and immunocompromised individuals may play in the antigenic evolution of SARS-CoV-2 (and other pathogens). Modelling of immunocompromised individuals during the COVID-19 pandemic has largely focused on their increased risk of mortality, rather than their potential importance for pathogen evolution [48–50]. Our study emphasises the need to consider both aspects. We also did not explicitly model within-host dynamics in the main text, instead approximating these dynamics following analysis of a separate within-host model in the appendix. The within-host model in the appendix demonstrated that immune pressure leads to diffusion through the antigenic space at a constant rate, therefore justifying our assumption of a constant rate of antigenic evolution in our primary model. This allowed us to assume infected individuals would substitute variant *i* with variant *i* + 1 at a constant rate, which mimicked typical within-host dynamics without the need for a fully nested model, which would be much more complex but would likely provide no additional insights to our simpler model.

We stress that while our results suggest that infected immunocompromised individuals may play a significant role in the antigenic evolution of SARS-CoV-2, we urge caution in how this message is interpreted and communicated. We urge particular caution with regards to the implications of our results for people who are immunocompromised. People may be immunocompromised for a variety of reasons, including uncontrolled HIV, undergoing treatment for cancer, or as a transplant recipient, and some conditions still wrongly attract stigma. Although Omicron was first detected in South Africa, which is estimated to have the highest HIV prevalence in the world (7.7 million people, with many infections uncontrolled [51]), this variant may have evolved in an individual without HIV and may have evolved elsewhere. Rather than stigmatising people who are immunocompromised, our results emphasise the need for global health equality and for better genomic surveillance, especially for immunocompromised people infected with SARS-CoV-2. Improving access to vaccines and treatments, especially in lower- and middle-income countries, and facilitating wider surveillance for new variants is crucial for limiting the emergence of new variants in the COVID-19 pandemic.

## Data Availability

Source code for the simulations is available in the Supplementary Material and at: https://github.com/ecoevotheory/Smith_and_Ashby_2022

https://github.com/ecoevotheory/Smith_and_Ashby_2022

## Appendix

### Within-host model

The model in the main text focuses on population-level dynamics and implicitly models within-host dynamics by assuming that: (1) immunocompetent and immunocompromised hosts differ in terms of their average infectious period; and (2) antigenic evolution occurs at a constant rate. Here, we consider the dynamics of a simple within-host model to justify the implicit within-host dynamics in our population-level model.

Let *V*_*i*_ be the viral abundance of variant *i* ∈ {1, …, *n*} within a single infected host and let *R*_*i*_ be the strength of the corresponding immune response. The virus grows exponentially with rate *r* in the absence of an immune response and decreases through the immune response at rate 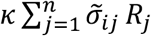, where *κ* is the per-capita rate of virus removal by the host immune system and 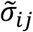 is the probability that an immune response for variant *j* causes cross-immunity to variant *i* such that

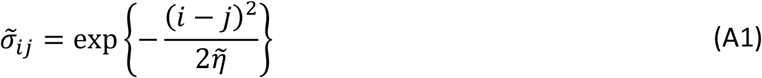

where 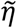 controls the breadth of cross-immunity between variants (similar to *η* in the main text). The virus also mutates to adjacent variants in the antigenic space with rate 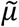. The immune response to variant *i* increases at per-capita rate *kqV*_*i*_ and decays with rate *d*. The parameter *q* controls the strength of host immune system such that larger values indicate an immune system that can respond well to infection (immunocompetent) and smaller values indicate a weaker immune response (immunocompromised).

As with the between-host model, we use the stochastic *τ*-leaping method [29] to simulate the within-host dynamics, corresponding to the following set of ODEs

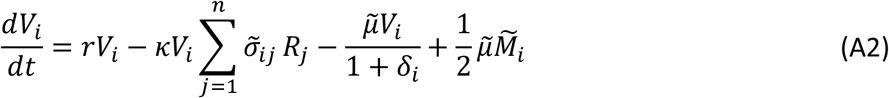

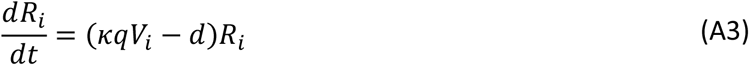

where 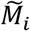 is the set of variants adjacent to *i* in the one-dimensional antigenic space (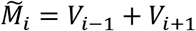 for *i* ∈ {2, …, *n* − 1}, with boundary conditions 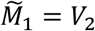 and 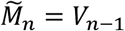), and *δ*_*i*_ = 1 if *i* ∈ {1, *n*} and is 0 otherwise to control the mutation rate at the boundaries.

When the host is immunocompetent (large *q*), the infection is rapidly cleared, with little within-host evolution (Fig. A1a). But when the host is immunocompromised (small *q*), the infection persists over much longer timescales, with immune pressure leading to successive selective sweeps as the virus diffuses through the antigenic space at a constant rate (Fig. A1b). If the mutation rate is faster in immunocompromised hosts (larger 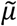), the coevolutionary dynamics of the virus and the immune response are simply accelerated (Fig. A1c compared with Fig. A1b). These results justify the simplifying assumptions in our population-level model regarding within-host dynamics, where we assume that there is a constant rate of antigenic evolution, which may differ between host types.

**Figure A1:**
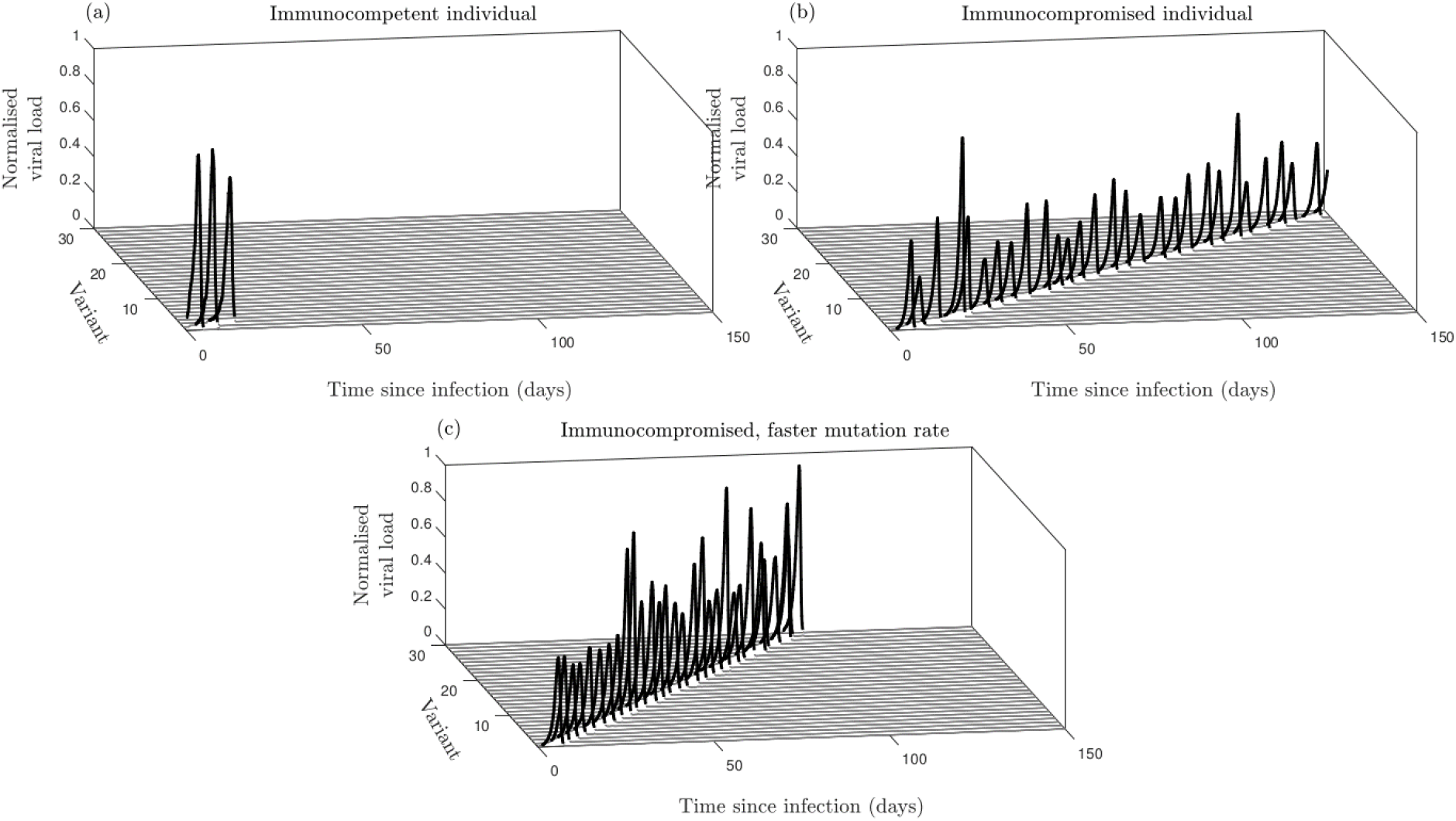
Within-host dynamics. *(a) Immunocompetent host* (*q* = 1.0), *(b) Immunocompromised host* (*q* = 0.1), *(c) immunocompromised host with faster mutation q* = 0.1, 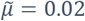). *Values of the viral load are normalised by the maximum value attained. All other parameters as in Table A4. Dynamics are shown for a single simulation*.

### Simulation algorithm

We simulate our within-host and population-level models using the *τ*-leaping method [29], which is an approximate stochastic simulation algorithm. We define the propensity functions 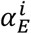 in Table A1, which give the rates of event type *E* for each variant index *i*. These propensity functions are then used to update the system synchronously at a time interval of one day using random numbers from the Poisson distribution 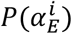. Source code for the simulations is available in the Supplementary Material and at https://github.com/ecoevotheory/Smith_and_Ashby_2022.

**Table A1:** The propensity functions for each of the event types in the population-level model.

| Event | Rate |
| --- | --- |
| Infection of host type $k \in \{H, C\}$<br>by variant $i$ | $\alpha_{\text{Inf}, k}^i = \beta_i S_k^i (I_H^i + I_S^i)$ |
| Gain of full immunity to variant $i$<br>due to infection by variant $j$ (host<br>type $k \in \{H, C\}$) | $\alpha_{\text{Imm}, j, k}^i = \beta_j \sigma_{ij} S_k^i (I_H^j + I_S^j)$ |
| Recovery by host type $k \in \{H, C\}$<br>from variant $i$ | $\alpha_{\text{Rec}, k}^i = \gamma_k I_k^i$ |
| Mutation from variant $i$ to variant<br>$i \pm 1$ for host type $k \in \{H, C\}$. | $\alpha_{\text{Mut}, k}^i = \frac{\mu_k}{1 + \delta_i} I_k^i$ |

**Table A2:** The propensity functions for the various event types for the within-host model.

| Event | Rate |
| --- | --- |
| Growth of variant $i$ | $\alpha_{\text{Grow}}^i = r V_i$ |
| Removal of variant $i$ by the<br>corresponding immune response | $\alpha_{\text{Rem}}^i = \kappa V_i R_i$ |
| Removal of variant $i$ by immune<br>response with index $j$ | $\alpha_{\text{Cross-rem}, j}^i = \kappa \tilde{\sigma}_{ij} V_i R_j$ |
| Mutation from variant $i$ to variant<br>$i \pm 1$ | $\alpha_{\text{Mut}}^i = \frac{\tilde{\mu}}{1 + \delta_i} V_i$ |
| Decay of immune response for<br>variant $i$ | $\alpha_{\text{Dec}}^i = d R_i$ |

**Table A3:**
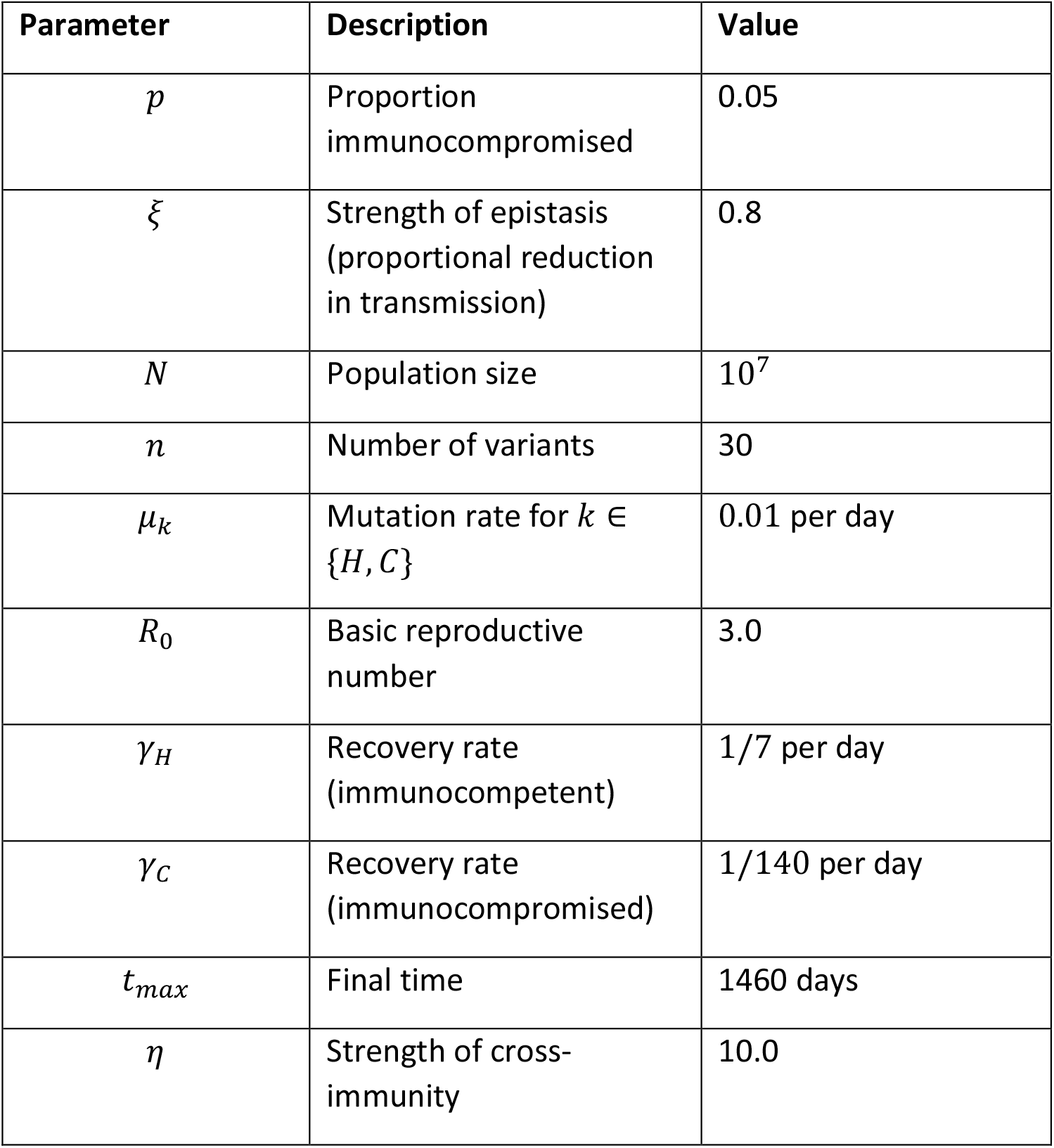
Default parameters for the between-host model.

**Table A4:**
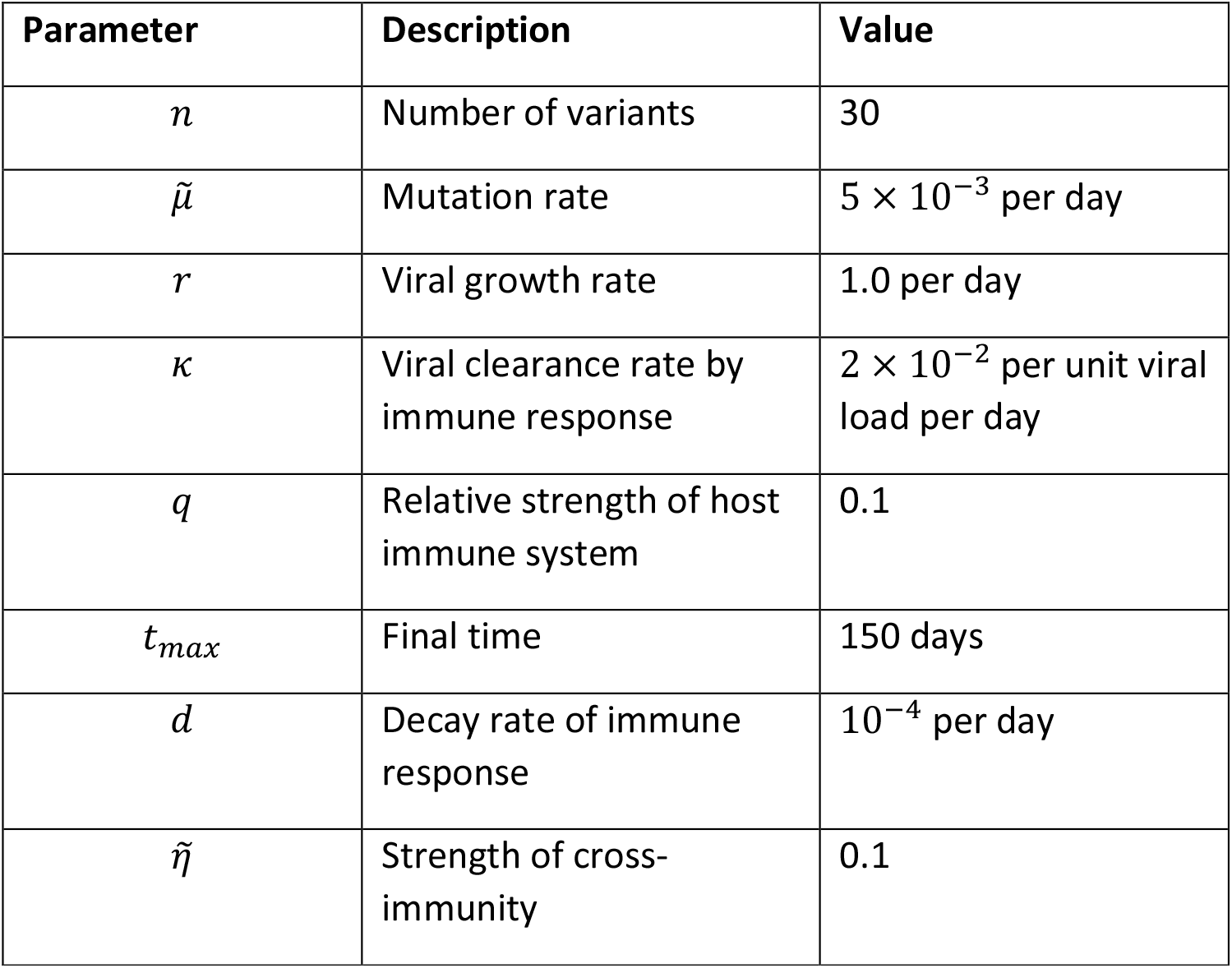
Default parameters for the within-host model.

## Acknowledgements

We thank Angus Buckling for helpful discussions. CAS is funded by the Natural Environment Research Council (NE/V003909/1). BA is funded by the Natural Environment Research Council (NE/N014979/1 and NE/V003909/1).

## Author contributions

BA conceived the study, CAS carried out the analysis, and both co-authored the manuscript.

